# Quantifying post-vaccination protective anti-SARS-CoV-2 IgG antibodies in blood and saliva with a fully automated, high throughput digital immunoassay

**DOI:** 10.1101/2022.01.21.22269165

**Authors:** Joseph M. Johnson, Syrena C. Fernandes, Danica L. Wuelfing, Aaron R. Baillargeon, Evan L. MacLure, Soyoon Hwang, Andrew J. Ball, Narayanaiah Cheedarla, Hans P. Verkerke, Sindhu Potlapalli, Kaleb Benjamin McLendon, Andrew Neish, William O’Sick, John D. Roback, David H. Wilson, Dawn Mattoon

## Abstract

**Background:** Antibodies induced by COVID-19 vaccination have been shown to wane over time. Current tests for assessing virus-neutralizing antibodies are complex and time-intensive. There is a need for a simple diagnostic test that measures levels of protective antibodies to help monitor immunity status.

**Method:** Using a commercially available FDA-authorized semi-quantitative SARS-CoV-2 IgG test, we monitored the duration of the immune response in dried blood microsamples (DBS) and saliva to vaccination by 3 different vaccines across prospective cohorts of 8 COVID-19 naïve and 29 COVID-19 recovered individuals over a six-month period. We correlated the results to a binding blockade assay validated to a live virus neutralization assay to validate the test for measurement of protective antibodies.

**Results:** The immune response characteristics between the two mRNA vaccines were similar over the 6-month period in both the COVID-19 naïve and recovered cohorts. IgG titers in DBS were generally 3-4 orders of magnitude higher than in saliva, and longitudinal profiles were highly correlated between the two matrices (R_m_ = 0.80). Median IgG concentrations post-vaccination declined to <10% neutralization capacity with all vaccines by six months.

**Conclusions:** The potential of a simple, fully automated high throughput anti-SARS-CoV-2 IgG test to quantitatively measure protective antibodies in samples collected remotely or at the point of care was demonstrated. The IgG immune response and protective immunity was shown to decline significantly by six months.

**Plain Language Summary:** In response to infection the immune system produces proteins called antibodies that recognize and bind to foreign invaders. Vaccines train the immune system to recognize and produce antibodies against specific invaders, such as SAR-CoV-2. Measurement of antibody levels in blood help monitor a person’s response to vaccination and have been shown to correlate with protection against disease, which wanes over time following vaccination. It is desirable to have an easy test that predicts protection against infection and measuring antibody levels may provide a solution, however different tests report results differently hindering the establishment of a cutoff for protected vs. not. We quantified antibody levels in saliva and dried blood microsamples (DBS) following vaccination using an automated semi-quantitative IgG test. By reporting concentration of antibodies, and if anchored to an international standard, this test could help establish a cutoff of protection that would be transferable across the multiple different test types. Furthermore, by measuring in saliva and DBS we demonstrate an easy path to at-home or point-of-care sample collection, which could allow wide-scale monitoring of immune protection against SARS-CoV-2.

## Introduction

Severe acute respiratory syndrome coronavirus 2 (SARS-CoV-2) is the cause of an ongoing pandemic that has infected more than 260 million people and caused more than five million deaths worldwide. The rapid development of safe and effective vaccines has enabled nearly eight billion vaccine doses to be administered, and it is estimated that approximately 43% of the global population has received a full initial vaccine regimen. Two-dose mRNA vaccine regimens elicit a generally robust humoral antibody response in previously healthy individuals, with a significant augmentation in circulating anti-spike protein IgG antibodies following administration of the second dose of either the BioNTech/Pfizer BNT162b2 vaccine or Moderna’s mRNA-1273 vaccine.^1,2^ Importantly, vaccine efficacy correlates with levels of neutralizing antibodies as measured by microneutralization testing,^3^ and levels of neutralizing antibodies correlate with total circulating IgG.^1^ Microneutralization assays require a high degree of biosafety due to handling of live active virus and are difficult to scale, making total IgG measurements attractive as a correlate of protection.

While the antibody response following SARS-CoV-2 vaccination is generally robust, it is important to understand the kinetics, amplitude, and durability of antibody responses at both the individual and population level to enable maintenance of protective immunity. Peak antibody titers have been reported six to ten weeks following administration of a two-dose mRNA vaccine, with significant variability observed in antibody response among vaccinated individuals.^1,4^ Attenuated antibody responses have been reported in immunocompromised individuals, including transplant recipients, suggesting these individuals remain vulnerable to infection following standard vaccination protocols.^5^ Conversely, higher peak antibody titers are reported in vaccinated patients with previous COVID-19 infection compared to COVID-19 naive patients.^4,6^ Studies evaluating antibody and antigen-specific memory B-cell responses in SARS-CoV-2 naive and recovered individuals suggest that responses are significantly augmented in individuals with previous COVID-19 infection, with no significant increase in circulating antibodies, neutralizing antibody titers, or antigen-specific memory B cells following the second dose of vaccine.^7^ Given global vaccine supply shortages and the potential for side effects from vaccination, it will be important to further characterize the optimal vaccine regimen for recovered individuals. Taken together, the variability in antibody response to COVID-19 vaccination between different populations suggests that the interval between initial vaccination and vaccine booster administration might benefit from tailoring to maintain protective immunity in healthy individuals, while avoiding unnecessary vaccine-induced side effects in those with a more durable protective immune response.

Longitudinal studies indicate the level of total IgG declines during the six months following vaccination, and neutralizing antibody titers decline rapidly for the first three months post vaccination.^2,8^ Studies modeling the longitudinal decay of neutralizing antibody titers following vaccination predict a significant loss in protection from SARS-CoV-2 infection and suggest that the duration of protective immunity is a function of initial vaccine efficacy as measured by neutralizing antibody titer.^3^ These results further underscore the need for quantitative correlates of protection.

The informed development and implementation of global public health policies aimed at pandemic management depend on accurate data on SARS-CoV-2 immunity at the population level. Achieving this requires the availability of simple, non-invasive tools to assess immunity that can be broadly deployed. Previous studies have demonstrated the potential for detection of SARS-CoV-2 antibodies in saliva that show strong correlation to serum IgG responses, with similar temporal kinetics.^9^ More recently, antibody response monitoring in saliva has been utilized as a surveillance tool to estimate viral transmission dynamics in young populations.^10^

Here we report the results from a six-month prospective longitudinal study in which we collected dried blood microsamples (DBS) and saliva from COVID-19 naive and recovered volunteers at defined intervals prior to, and following vaccination. The saliva and dried blood samples were tested for SARS-CoV-2 anti-spike IgG in a high sensitivity semi-quantitative antibody test.^11^ We observed robust anti-spike IgG responses in all subjects receiving mRNA vaccines and higher peak antibody response in subjects with prior COVID-19 infection. COVID-19 naive and recovered subjects exhibited declining levels of circulating anti-spike IgG with similar kinetics. Antibody levels in DBS correlated strongly with those in saliva with similar temporal kinetics. We also found that total anti-spike IgG levels in DBS correlated strongly with the ability to block binding of spike protein to the ACE2 receptor and to levels of neutralizing antibodies. This work describes a path for high throughput quantitative immunity assessment and monitoring in readily obtained samples that can be collected remotely or at the point of care.

## Methods

### Apparatus

Anti-SARS-CoV-2 IgG and ACE2 receptor binding blockade assays were run on the Quanterix Simoa HDX Analyzer, which is a fully automated immunoassay analyzer utilizing Simoa (single molecule array) technology for isolation and counting of single molecules. In brief, Simoa digitizes bead-based ELISAs whereby the diffusion of fluorescent reporter molecules at the signal step is constrained to 40-femtoliter wells in a microarray of 216,000 microwells. By restricting diffusion to such a small volume, fluorophores generated by a single enzyme label can be detected in the array in 30 seconds, and the large number of microwells enables the measurement of many wells simultaneously for robust statistics. The HDX pipettes sample directly from sample tubes or 96-well plates and processes assays and data reduction with a steady state throughput of 66 tests/hour. Details of the technology and instrument are given elsewhere.^12,13^

### Anti-SARS-CoV-2 IgG assay and research protocol

Anti-SARS-CoV-2 IgG measurements were made with the Simoa^®^ Semi-Quantitative SARS-CoV-2 IgG Antibody Test.^11^ This assay is a 3-step paramagnetic microbead-based sandwich ELISA. In the first step, sample is drawn from a sample tube or microtiter plate by the instrument pipettor and mixed with COVID-19 spike protein coated paramagnetic capture beads in a reaction cuvette. IgG antibodies in the sample that are specific to COVID-19 spike protein are bound by the capture beads. After an incubation, capture beads are collected with a magnet, and washed. Biotinylated anti-human IgG detector antibodies are then mixed with the capture beads, and the detector antibodies bind to the captured sample IgG. Following a second wash, a conjugate of streptavidin-ß-galactosidase (SBG) is mixed with the capture beads. SBG binds to the biotinylated detector antibodies, resulting in enzyme labeling of captured sample IgG. After a third wash, the capture beads are resuspended in a resorufin ß-D-galactopyranoside (RGP) substrate solution. Digital processing occurs when beads are transferred to the Simoa array disc.^14^ Individual capture beads are then sealed within microwells in the array. If IgG antibodies have been captured and labeled, the ß-galactosidase hydrolyzes the RGP substrate into a fluorescent product that provides the signal for measurement. The concentration of IgG in unknown samples is interpolated from a standard curve obtained by 4-parameter logistical (4PL) regression fitting. Total time to first result is 60 minutes.

The anti-SARS-CoV-2 IgG assay is calibrated with a chimeric human/mouse anti-SARS-CoV-2 IgG. This is a chimeric monoclonal antibody combining the constant domains of the human IgG1 molecule with mouse variable regions. The variable region was obtained from a mouse immunized with purified, recombinant SARS-CoV spike S1 protein, and the antibody was produced using recombinant antibody technology. The immunogen for this molecule (spike S1 protein) is the same as the antigen coated on the solid phase of the assay capture microbeads. Analytical studies to support the validity of the chimeric IgG calibrators included a comparison of the dilution properties of native positive samples relative to the calibrators; good parallelism was obtained indicating that the binding interactions between assay reagents and captured chimeric anti-SARS-CoV-2 IgG provide a good mimic for the interactions against captured patient sample anti-SARS-CoV-2 IgG (data not shown). A weight concentration was assigned to the purified chimeric IgG, from which a set of 8 assay calibrators of known concentration are prepared and from which signals from patient samples can be converted into μg/mL. Note that because these standards are not currently harmonized to the recently available First WHO International Standard for anti-SARS-CoV-2 immunoglobulin (NIBSC 20/136, South Mimms, England), the assay is referred to as “semi-quantitative.”

### Anti-SARS-CoV-2 IgG positive / negative cutoff

For research purposes, a positive/negative cutoff of 0.77 µg/ml previously established by the assay kit manufacturer for serum and plasma was applied to DBS in view of the large (1000-fold) dilution for all three sample types. A positive/negative cutoff was not established for saliva samples.

### SARS-CoV-2 ACE2 receptor binding blockade of ACE-2 binding assay and protocol

Levels of SARS-CoV-2 neutralizing antibodies were estimated using an in-house developed homebrew assay on the Simoa HDX analyzer (ref Emory companion manuscript submitted in parallel). Paramagnetic beads were covalently conjugated with a soluble trimeric spike ectodomain based on the sequence of Wuh-1 SARS-CoV-2. Samples eluted as described for the IgG assay were diluted to a final sample dilution of 1:50 before incubation with target beads in a 3 step HDX assay, using a biotinylated ACE-2 IgFc chimera as the detector. Blockade of ACE-2 binding was estimated for each sample in duplicate as a % inhibition normalized to the maximal ACE-2 binding signal observed in buffer controls.

### Sample collection and preparation

This study was IRB approved (Advarra) and all participants provided written informed consent. Samples were blinded before testing.

#### Dried Blood Microsamples (DBS)

DBS fingerstick samples were collected using Neoteryx^®^ Mitra® 20μL micro sampling kit (Item # 200307, Neoteryx, Torrance, CA) according to the manufacturer’s instructions for use. Samples collected with this device are stable for up to two weeks at room temperature, after which they were kept at -20°C. To extract sample from the Mitra microsampler tips, 200 μL of anti-SARS-CoV-2 IgG assay Sample Diluent was pipetted into the wells of a 96 deep-well plate (1 mL/well). Each microsampler tip was placed into a well containing Sample Diluent. The tips were eluted overnight (16-18 hours) with shaking at 400 rpm at 2-8°C. Extraction results in a nominal 10-fold dilution (20 uL to 200 uL). DBS extracts were then further dilution 100-fold with Sample Diluent, resulting in a total dilution of 1000-fold. For testing, samples were pipetted into wells of the Simoa 96 well assay plate (230 μL for duplicate results). Results were multiplied by the dilution factor of 1000 before reporting.

#### Saliva samples

Saliva samples were obtained from repeated spiting into a sterile, leak-proof screw cap container (50mL centrifuge tubes, Thermo Fisher Scientific, USA) until approximately 3 ml of liquid was collected (excluding bubbles). Samples were stored at -20°C prior to testing. After thawing, a clear supernatant was separated from solids and viscous material, transferred to a separate tube, vortexed vigorously and then sub-aliquoted into 500 ul vials. Aliquots could be tested the same day or stored at - 20°C. Before testing, aliquots were centrifuged at 3000 x g for 10 minutes, a clear supernatant was removed which was easy to pipette and not viscous. Saliva supernatants were diluted either 10 or 20-fold with sample diluent and pipetted into the wells of the Simoa 96 well assay plate (230 μL for duplicate results). Samples with IgG levels above the calibration curve were retested using frozen aliquots diluted 50-fold. Batch assays were initiated following the HDX user guide procedure for inserting the 96 well assay plates into the instrument and initiating the run. Results were multiplied by the applicable dilution factor before reporting.

### Study Cohorts

Matching saliva and DBS were collected from 13 donors pre- and post-vaccination out to 6 months, with frequent sampling during weeks one and two; an additional 2 donors donated only saliva (cohort 1). DBS only was collected from 24 donors pre- and post-vaccination out to 6 months with less frequent sampling (cohort 2). The details of donor numbers and collection time points for both cohorts are shown in Table 1 and Figure 1. Donors were determined as having a prior-COVID infection through self-attestation based upon positive results of either PCR or antigen tests except for one donor who was presumed to have been prior-exposed based upon a rapid rise in IgG levels <1 week after first vaccination. Complete donor details are available in Supplementary Information Table S1.

**Table 1.**
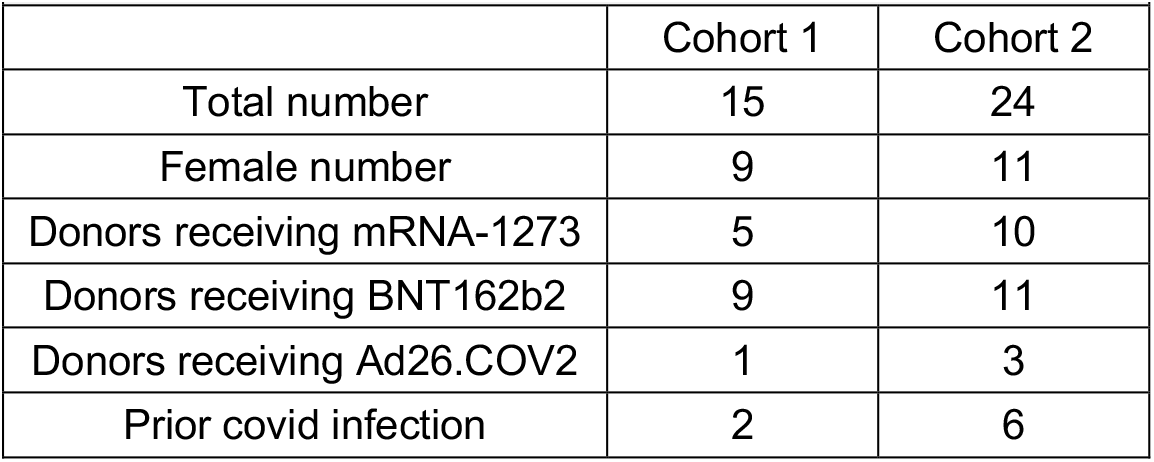
Donor details of cohort 1 and 2.

| Table 1. Donor details of cohort 1 and 2. |  |  |
| --- | --- | --- |
|  | Cohort 1 | Cohort 2 |
| Total number | 15 | 24 |
| Female number | 9 | 11 |
| Donors receiving mRNA-1273 | 5 | 10 |
| Donors receiving BNT162b2 | 9 | 11 |
| Donors receiving Ad26.COV2 | 1 | 3 |
| Prior covid infection | 2 | 6 |

**Fig. 1.**
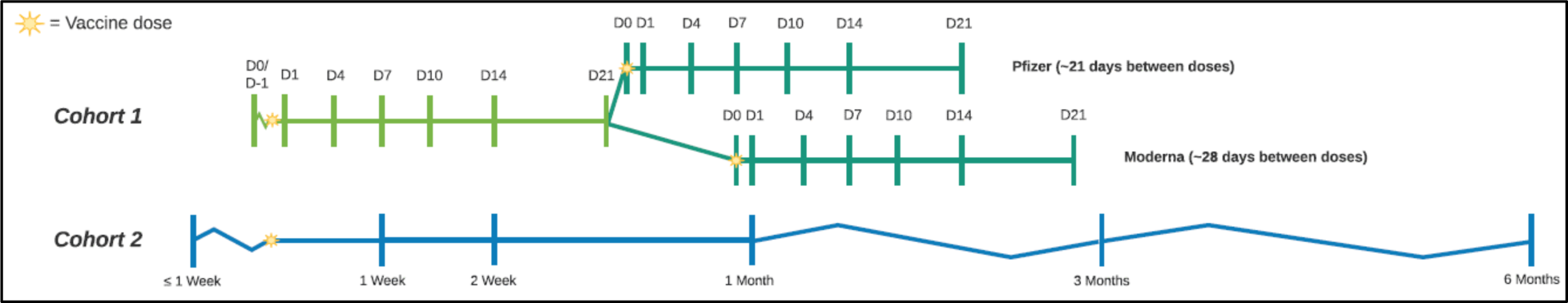
Timepoints of sample collection for cohort 1 and 2.

### Statistical Analysis

Statistical analyses were performed using Graphpad prism (version 8.4.0 (671), Microsoft Excel (16.0.13530.20132), or R studio, R v.4.0.3 (package rmcorr).^15^ Sample size for each cohort was not determined based on statistical hypothesis testing; all donors meeting study criteria were included. Descriptive statistics, including median and interquartile range, were reported for continuous data with a skewed distribution. Categorical measures, such as percent of donors seroconverted, were reported as a percentage along with a Clopper-Pearson 95% confidence interval. Repeated measures correlation was used to assess the within-participant association of DBS and saliva IgG levels over time in Cohort 1, using longitudinal measurements spanning from pre-vaccination through 67 days post-vaccination.^16^ A Mann-Whitney test was conducted for nonparametric comparison between COVID naive and previously infected donors in Cohort 2 at each timepoint, with a Bonferroni correction for multiple hypotheses. Percent inhibition as a function of IgG concentration and duration of protection based on peak IgG levels were fit with 4PL regression in the Graphpad Prism (Fig 3 and S3) of the form: Y=Bottom + (X^Hillslope)*(Top-Bottom)/(X^HillSlope + EC50^HillSlope). Decay curves of IgG levels were fit with one-phase exponential decays in Graphpad Prism (Fig 4e and Supplementary Figure S2) of the form: Y=(Y0 - Plateau)*exp(-K*X) + Plateau.

## Results

### Longitudinal IgG concentrations in COVID naive and recovered individuals

We measured pre- and post-vaccination anti-SARS-CoV-2 IgG profiles over a six-month period across three different vaccines for COVID-naïve (Fig. 2a) and COVID-prior-infected donors (Fig. 2b). Two weeks after the initial dose 100% of COVID-naïve donors receiving mRNA vaccines had seroconverted, with peak levels observed at one month. Figure 2a underestimates the max response of donors receiving mRNA-1273 because the 2^nd^ vaccination occurs simultaneously with the 1-month sample collection, contrasting with BNT162b2 vaccinated who received the 2^nd^ dose at 21 days. The results demonstrate variability in the amplitude of individual antibody response to vaccination, consistent with previous studies.^17,18^. For both mRNA vaccines, median antibody concentrations show measurable decline by the six-month time point following initial vaccination. In this small cohort (N=2), antibody levels in subjects receiving the Ad26.COV2.S adenoviral vector vaccine rose with slower kinetics and achieved lower peak concentrations.

**Fig. 2.**
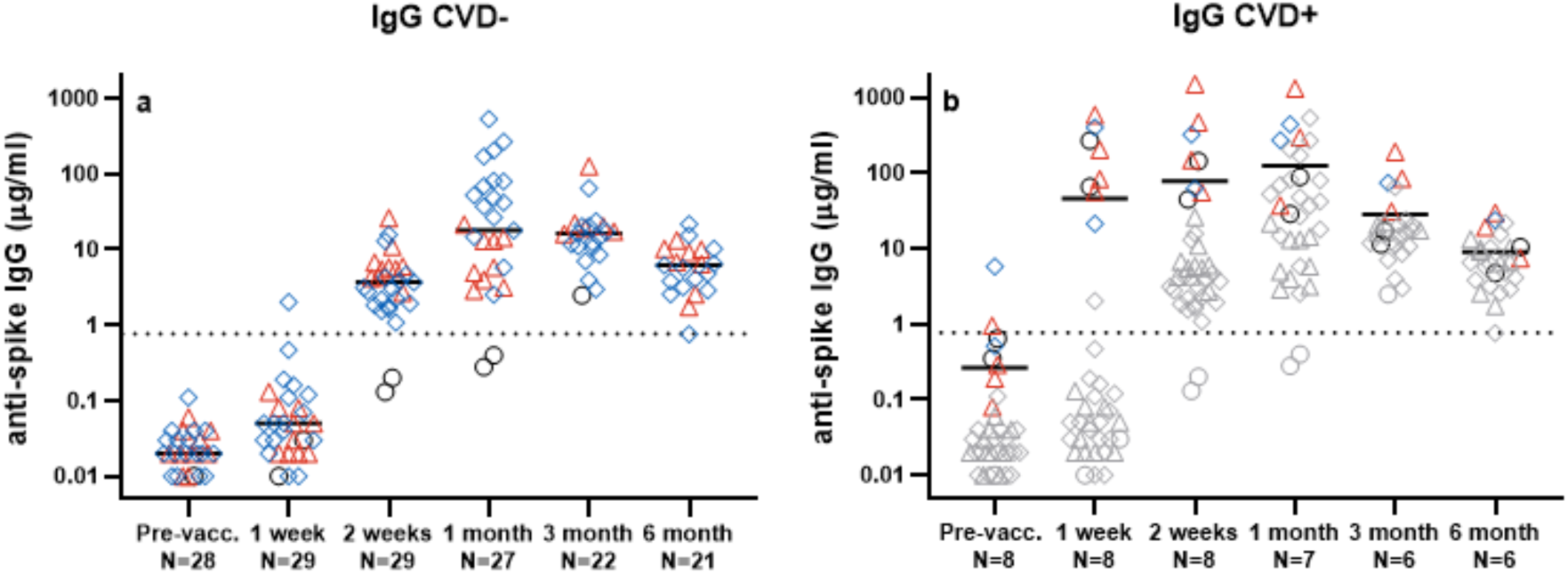
SARS-CoV-2 anti-spike IgG in dried blood microsamples pre- and post-vaccination in COVID-19 naive and recovered individuals. **a)** Covid-naive donors. **b)** Donors previously diagnosed with COVID-19 are highlighted. COVID-19-naive donors (previous panel) are shown in grey. Symbols: blue diamonds, Pfizer/BioNTech (BNT162b2); red triangles, Moderna (mRNA-1273); black circles, Janssen/J&J (NJ-78436735). Solid black lines denote the median value; dotted black line the positive / negative cutoff for the Simoa SARS-CoV-2 Semi-Quantitative Antibody Test. Data is binned from the time of first vaccination dose. N= number of individual donors.

**Fig. 3.**
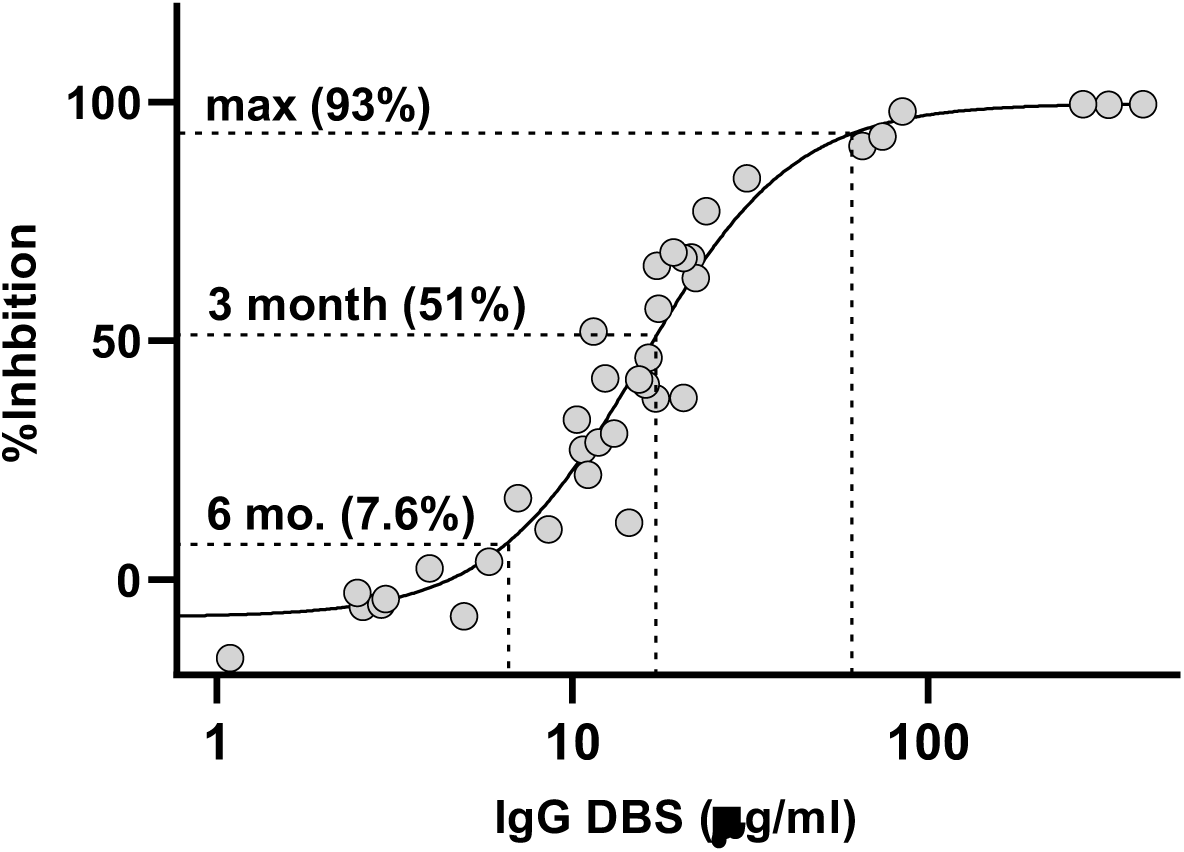
Correlation of SARS-CoV-2 anti-spike IgG concentration and anti-spike neutralization activity. Pseudo-neutralization (% inhibition) as a function of IgG concentration for DBS samples (50 samples from 29 individual donors). Median IgG concentrations (for COVID naïve and prior-infected) at 3 month (28 donors) and 6 months (27 donors) are highlighted along with the corresponding % inhibition based upon four parameter logistic regression. The maximum IgG concentration observed for the longitudinal cohort is also denoted (median over 11 donors, only COVID naïve).

**Fig. 4.**
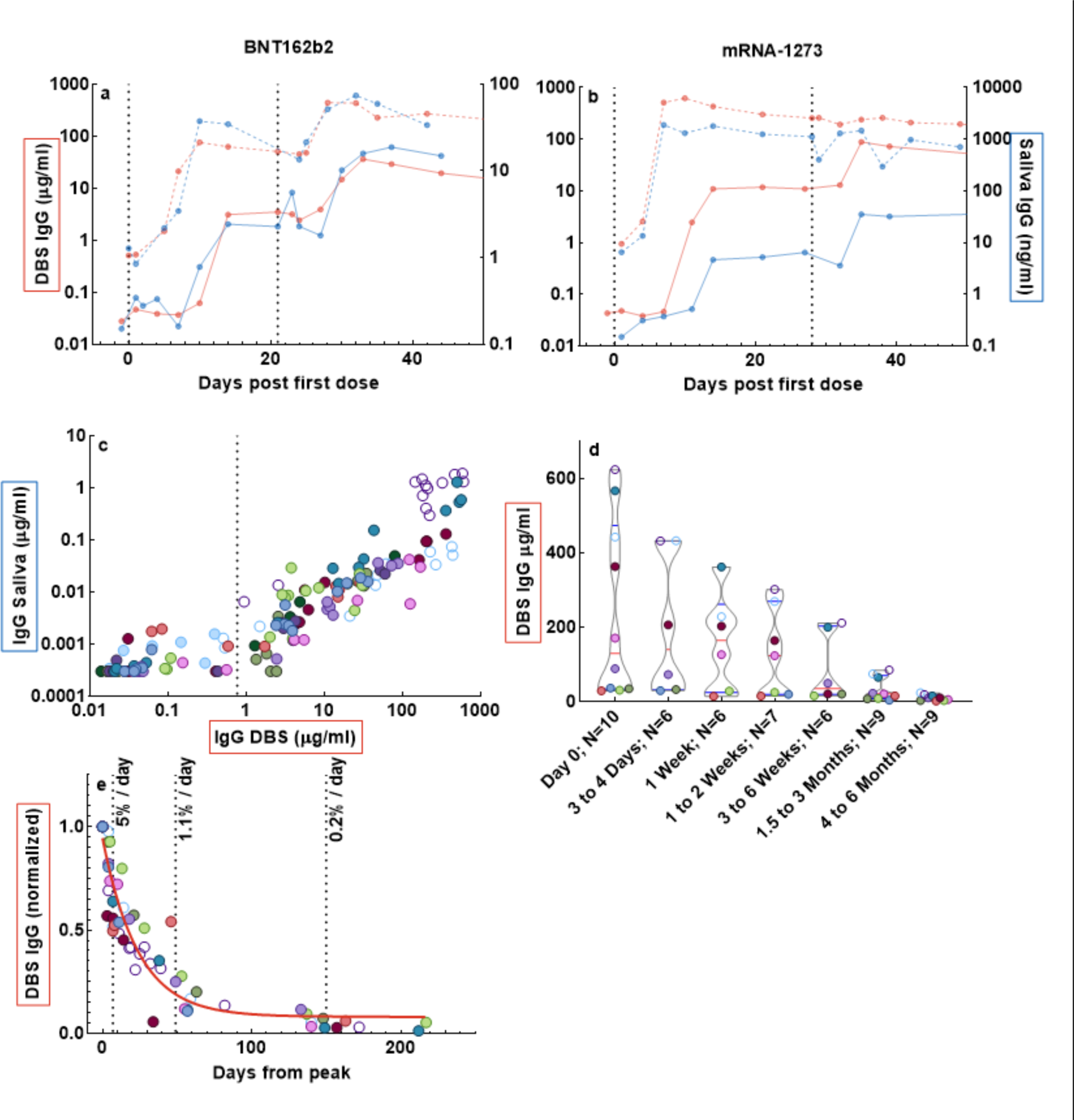
Temporal kinetics of antibody response in matched dried blood microsamples and saliva. SARS-CoV-2 anti-spike IgG in saliva strongly correlates with IgG in DBS. **a)** Representative IgG-longitudinal traces from BNT162b2 and **b)** mRNA-1273 inoculated donors are shown for matched DBS and saliva. The profiles indicate days from first dose of DBS (red, left y-axis) and saliva (blue, right y-axis) from naive (solid) or previously infected donors (dashed). The 1^st^ and 2^nd^ doses are shown as vertical dotted lines. **c)** IgG levels in longitudinal, matched DBS/saliva samples color-coded by donor (n=152 data points from n=13 donors). Open symbols indicate previously infected donors; the positive /negative cutoff in DBS is indicated with a vertical dotted line. **d)** Concentration of IgG from multiple donors binned by days from peak; donor number (N) is indicated in the x-axis label; median (red lines) and interquartile ranges (blue lines) are indicated for each time bin. **e)** The normalized response from peak is plotted as a function of days from peak (n = 61 data points, N = 10 donors). The mean decay rate (% / day) is calculated at 1 week (N = 10 donors), 7 weeks (N=10) and 5 months (N=9) post-peak from time-binned data (dotted vertical lines).The solid red line shows a fitted one-phase exponential decay.

As shown in Fig. 2b, 100% of donors with prior COVID-19 infection achieved seroconversion within 1 week; IgG levels rose faster and attained a higher peak than COVID-19 naive subjects. A Mann-Whitney nonparametric comparison with a Bonferroni correction for multiple hypotheses demonstrated significantly elevated IgG levels in previously infected donors at the pre-vaccination (p < 0.001), 1-week (p < 0.001), 2 weeks (p < 0.001) and 1-month (p < 0.019) timepoints. However, by 3 months post vaccination the difference in IgG levels was not statistically significant between previously infected and COIVD-naïve donors (p=0.199), and remained so at 6 months post-vaccination (p=0.130), suggesting vaccinated subjects are likely to receive similar durability of protective immunity independent of previous infection status. The limited number of Ad26.COV2.S recipients with prior COVID-19 infection (N=2) showed similar magnitude and duration of response as the mRNA vaccine recipients, raising the possibility that a multi-dose schedule of Ad26.COV2.S could achieve similar antibody titers. Quantification of the data in Figure 2 is shown in Table 2.

**Table 2.** Characteristics of IgG levels in donors over 6-month timepoint collection. Units are in µg/mL.

| **Table 2. Characteristics of IgG levels in donors over 6-month timepoint collection. Units are in $\mu\text{g/mL}$.** |  |  |  |  |  |  |  |  |  |
| --- | --- | --- | --- | --- | --- | --- | --- | --- | --- |
| **Covid Naïve** |  |  |  |  |  |  |  |  |  |
| Timepoint (first dose) | Percent > cutoff | 95% CI | Min. | 1st Quartile | Median | Mean | 3rd Quartile | Max. | N |
| Pre-Vacc. | 0% | 0%, 12% | 0.01 | 0.02 | 0.02 | 0.03 | 0.03 | 0.11 | 28 |
| 1 Week | 3% | 0%, 18% | 0.01 | 0.02 | 0.05 | 0.14 | 0.08 | 2.03 | 29 |
| 2 Weeks | 93%* | 77%, 99% | 0.13 | 1.94 | 3.68 | 5.06 | 5.58 | 26.09 | 29 |
| 1 Month | 93%* | 76%, 99% | 0.28 | 5.38 | 17.89 | 64.69 | 61.01 | 537.37 | 27 |
| 3 Months | 100% | 85%, 100% | 2.49 | 10.80 | 16.22 | 21.42 | 20.49 | 127.11 | 22 |
| 6 Months | 95% | 76%, 100% | 0.76 | 3.19 | 6.20 | 7.06 | 10.07 | 21.84 | 21 |
| **Previously Infected** |  |  |  |  |  |  |  |  |  |
| Timepoint (first dose) | Percent > cutoff | 95% CI | Min. | 1st Quartile | Median | Mean | 3rd Quartile | Max. | N |
| Pre-Vacc. | 25% | 3%, 65% | 0.08 | 0.26 | 0.43 | 1.10 | 0.71 | 5.82 | 8 |
| 1 Week | 100% | 63%, 100% | 21.28 | 63.59 | 143.76 | 211.62 | 303.05 | 589.29 | 8 |
| 2 Weeks | 100% | 63%, 100% | 44.71 | 60.88 | 146.89 | 343.92 | 359.11 | 1502.38 | 8 |
| 1 Month | 100% | 59%, 100% | 28.70 | 62.99 | 272.72 | 354.61 | 369.56 | 1315.73 | 7 |
| 3 Months | 100% | 54%, 100% | 11.46 | 20.67 | 52.65 | 68.38 | 82.11 | 191.60 | 6 |
| 6 Months | 100% | 54%, 100% | 4.74 | 8.22 | 14.87 | 15.92 | 22.64 | 29.79 | 6 |

\*100% seroconversion observed for mRNA vaccine recipients.

### Correlation of anti-SARS-CoV-2 IgG assay with protective immunity

To evaluate the significance the measured IgG concentrations in terms of protective immunity, we compared the assay readout to a validated binding blockade assay which measures the inhibition of the interaction between ACE2 and spike protein by protective antibodies, not necessarily limited to IgG class. The binding blockade assay is outlined in the Methods section and is described in detail elsewhere (ref Emory companion manuscript submitted in parallel It has been shown to correlate strongly with anti-SARS-CoV-2 neutralization activity as measured with a live virus focus reduction neutralization test, considered to be a gold standard, as has been observed in other studies.^19^ Fifty DBS samples collected from 29 vaccinated subjects collected longitudinally over a six-month period were tested in the binding blockade assay, and percent inhibition of binding as a function of anti-spike IgG concentration measured by the anti-SARS-CoV-2 IgG test was determined (Fig. 3). Median IgG levels in the interval of peak antibody concentration correspond to 93% inhibition of binding between ACE2 receptor and spike protein, with inhibition of binding declining to 51% at the three-month median IgG concentration, and inhibition further declining to just 7.6% at the six-month median IgG concentration. These data suggest protective immunity may be significantly attenuated within six months of vaccination, underscoring the importance of boosting immune response through a semi-annual vaccination regimen as currently recommended by public health guidelines.^20^

### Anti-SARS-CoV-2 IgG in matched DBS and saliva

To explore the ability to quantify anti-SARS-CoV-2 IgG in saliva, matched dried blood microsamples and neat saliva samples were self-collected longitudinally from a subset of 13 individual donors prior to and over a period of 6 months following vaccine administration. These samples were collected at a relatively higher frequency near the vaccination event, to quantify the rate of vaccination response at higher temporal resolution. We show representative individual data in Fig. 4a and b and individual traces for all 15 longitudinal donors (including 2 saliva only donors) are in Supplementary Fig. S1. Salivary antibody concentrations are higher in COVID-19 recovered individuals prior to vaccination and rise with accelerated kinetics relative to COVID-19 naive individuals, mirroring the results observed in dried blood samples.

By sampling at a higher frequency, we were more accurately able to define the peak IgG levels in both DBS and saliva, as shown in Table 3. Levels of anti-SARS-CoV-2 spike protein antibodies are generally 3-4 orders of magnitude lower in saliva samples relative to dried blood samples (average = 2051, range = 254 to 5991), however the high sensitivity of the Simoa digital immunoassay enables robust detection in both matrices. We find DBS peak levels 2 to 3 times higher than in DBS sampled less frequently (compare to Table 2). DBS and saliva samples demonstrate excellent concordance (Fig. 4c), particularly above the clinical cutoff, although levels in saliva appear more variable than those in DBS. A repeated measures analysis was applied to the COVID-19 naïve samples, yielding a strong correlation of 0.80 between DBS and saliva (R_m_=0.80, 95%CI = (0.71,0.86), p-value < 0.001).

**Table 3.** Characteristics of IgG levels in a subset of 13 donors for both DBS and saliva, sampled with higher frequency near vaccination.

|  |  |  |  |
| --- | --- | --- | --- |
| Peak DBS (µg/mL) |  |  |  |
| Infection Status | Median | 1st Quartile | 3rd Quartile |
| Covid Naïve | 60.1 | 32.8 | 129.6 |
| Previously Infected | 532.8 | 487.4 | 578.2 |
| Peak Saliva (ng/mL) |  |  |  |
| Infection Status | Median | 1st Quartile | 3rd Quartile |
| Covid Naïve | 29.0 | 20.7 | 45.6 |
| Previously Infected | 982.5 | 528.2 | 1436.9 |

We quantified the kinetics of the anti-SARS-CoV-2 IgG response in DBS following vaccination (Table 4). We found a median delay of 10 days from first dose to the first quantifiable increase in IgG levels and 13 days until seroconversion (> 0.77 µg/mL). IgG levels then tend to plateau until the 2^nd^ dose is given, after which a second increase occurs. As expected, antibody levels respond faster to the 2^nd^ dose, with a delay of only 4 days from dose to first increase, and 10 days from 2^nd^ dose to the peak IgG level. Saliva follows similar kinetics, although response to the 2^nd^ vaccination is slightly delayed compared to DBS (Table 4). IgG levels varied more widely in saliva than DBS, with peaks often occurring near the time of vaccination, but then quickly decaying again (Supplementary figure S1).

**Table 4.** Kinetic characterization of antibody levels following vaccination.

|  |  |  |
| --- | --- | --- |
| **Temporal Kinetics of IgG Increase in COVID-naïve donors (N= donor number)** | **Median (1<sup>st</sup> quartile; 3<sup>rd</sup> quartile)** |  |
|  | DBS | Saliva |
| Days from 1<sup>st</sup> dose to increase ($N_{\text{DBS}}=11$; $N_{\text{Saliva}}=13$) | 10 (10,10.5) | 10 (10,14) |
| Days from 1<sup>st</sup> dose to seroconversion ($N_{\text{DBS}}=10$) | 13 (10.25,14) | NA |
| Days from 2<sup>nd</sup> dose to increase ($N_{\text{DBS}}=9$; $N_{\text{Saliva}}=13$) | 4 (4,4) | 7 (4,7) |
| Days from 2<sup>nd</sup> dose to peak ($N_{\text{DBS}}=7$; $N_{\text{Saliva}}=9$) | 10 (8.5,10) | 14 (10, 15) |

We also quantified the decay kinetics of antibody levels in DBS after the peak response. Despite a wide distribution of peak concentrations, IgG levels in all donors collapse to similarly low values by 4 to 6 months post-peak, including those with prior infections (Fig. 4d), confirming the lack of significant difference between covid-naïve and covid-prior-infected 3 to 6 months post-vaccination we observed (Fig. 2). In Fig 4e we show IgG decay kinetics for all donors normalized to their peak response. From this data we determined the rate of decay for individual donors in three time-bins: decay within the first week post-peak; decay between week 1 to approximately 2 months; decay between approximately 2 months to 5 months. IgG levels decrease rapidly within the first week (5%/day) and then continuously more slowly out to 2 months (1.1%/day) and 5 months (0.2%/day). The normalized decay of all donors fit to a one-phase exponential decay with R^2^ = 0.89 and predicting a half-life of 16.4 days (95% CI 12.3 to 21.9 days), which aligns well with our observation of IgG levels decaying most quickly right after the peak response. Exponential decay fits to individual donors showed similar trends as shown in supplementary figure S2, with a median half-life of 17.9 days (14.7, 23.5 interquartile) across donors.

Given that IgG levels decay exponentially at a similar rate for all donors (Fig. 4e), peak IgG concentrations will have a significant, though non-linear, impact on durability of humoral protection. If we assume that 50% blocking of binding, corresponding to 15.7 µg/ml (Fig. 3), results in robust humoral protection, then we may predict the length of time donors are protected based upon their peak IgG levels and the exponential in fit in Fig. 4e. We display this data in Table 5 and in supplementary Fig. S3).

**Table 5.** Duration of humoral protection based upon peak IgG levels.

| Table 5. Duration of humoral protection based upon peak IgG levels |  |
| --- | --- |
| Peak IgG | Decay time to 50% inhibition |
| 150 $\mu\text{g/ml}$ | 86 days |
| 100 $\mu\text{g/ml}$ | 58 days |
| 50 $\mu\text{g/ml}$ | 32 days |
| 30 $\mu\text{g/ml}$ | 16 days |

## Discussion

This work adds to our understanding of the post-vaccination antibody response to SARS-CoV-2 in several ways. First, we employed high frequency sampling to characterize the IgG response profiles of the two mRNA vaccines in more detail than in previous recent reports. ^7,8,21–24^ After the initial dose of mRNA vaccine, we find 100% seroconversion in 2 weeks among COVID-naïve mRNA vaccine recipients, and in only one week among COVID-experienced recipients. The median IgG titer increase among COVID experienced participants was over two orders of magnitude in only a week, while the titer increase was much slower with COVID naïve participants. As reported by Goel *et al*.^7^ maximal titers were obtained in both cohorts by one month, at or near the timing of the second dose. Due to the one-week timing difference between the Pfizer and Moderna second doses, a direct comparison of maximal second dose titers achieved by the two vaccines was not possible. While one week may seem like a small difference, it is significant given the dynamic properties of IgG titers seen in these data for both vaccines. At maximal titers, the median anti-spike IgG level for COVID experienced participants was ∼6-fold higher than for COVID naïve participants, aligning with the results of Kramer *et al*. ^6^ After the second dose, BNT162b2-induced IgG titers increased by approximately 10-fold in the COVID naive participants, while there was no discernable titer increase among the COVID experienced cohort. These results add to other data ^6,21–23,25^ supporting the suggestion that a second dose of mRNA vaccine may be unnecessary for COVID experienced individuals.

Second, we carefully characterized the post-vaccination IgG decay kinetics and directly compared this response to the decay of neutralizing antibodies. Consistent with other reports ^8,25,26^ we found a significant decay in post-vaccination IgG over 6 months. Surprisingly, this IgG decay corresponded to a loss of neutralizing activity as estimated by ACE-2 binding inhibition from 93% to only 7.8% by month 6. A rapid decay of neutralizing antibodies and IgG has been reported others, ^26–28^ and our data are consistent with those of Canaday *et al*. ^26^ who found an 83% decline in post vaccination IgG, with 70% of study participants exhibiting decayed neutralization titers to undetectable levels by pseudo virus assay by month 6. The IgG decay profile we found exhibited a non-linear response, initially declining 5.5%/day within the first week of the peak and then slowing to a 0.2%/day decay by months 4 to 6. The non-linear decay in IgG titer we found is at odds with Levin *et al*.^8^ where a linear drop in titer over six months was described. A possible explanation for this difference is that the Simoa IgG assay detects full anti-spike activity, while the assay used by Levin *et al*. was an anti-RBD assay. Since the mRNA vaccines produce neutralizing antibodies to both RBD and non-RBD spike epitopes,^29^ it is possible that the fuller antibody response detected by an anti-spike assay more closely reflects a non-linear decay of neutralizing antibodies. This observation also suggests that antibodies directed against non-RBD epitopes contribute to the in overall efficacy of the mRNA vaccines studied here. This is supported by the fact that efficacious neutralizing antibodies to non-RBD epitopes on the full spike protein have been described.^30^

Third, employing digital immunoassay technology, we were able to quantitatively characterize the longitudinal post-vaccination salivary IgG response with direct measurements in whole saliva and compare this response to humoral IgG. Due in part to the low concentration of salivary antibodies (generally below the detection limits of conventional ELISAs), there are limited quantitative data available on salivary anti-SARS-CoV-2 IgG. In line with another recent report comparing saliva to serum,^31^ we found vaccine-induced anti S-protein IgG in saliva 3 to 4 logs lower than capillary blood IgG. Dynamics of IgG induction were remarkably correlated between capillary blood and saliva (R_m_ = 0.80) across the first month post vaccination, suggesting a simple saliva test could be used to assess vaccination immunity status. As noted by Sheikh-Mohamed *et al*.^32^ salivary IgG titer declined precipitously over six months concomitant with a decline observed in humoral IgG. Antibodies in saliva are known to be dominated by secretory IgA (SIgA) and IgG. While SIgA is produced as dimeric IgA by plasma cells in salivary gland stroma, IgG in saliva largely originates from blood circulation by leakage mainly via gingival crevicular epithelium.^33^ The strong correlation between vaccine induced saliva and capillary blood-derived IgG adds to previous data^34–36^ suggesting this passive conduit may be a means to reliably monitor anti-SARS-CoV-2 IgG status and could be a viable alternative to venous and finger stick sample collection which may not be ideal for broad surveillance, including sampling from children.^34^ Digital immunoassay technology provides the analytical capability to precisely quantify salivary IgG directly in whole saliva with high throughput.

Vaccination programs seek to deliver individual acquired immunity, decreasing the probability of transmission to a level where viral propagation within a community is significantly diminished or even eliminated. The point at which the fraction of susceptible individuals falls below the threshold required for transmission is known as herd immunity.^37^ Communities achieving a vaccinated population above this threshold enables susceptible individuals who cannot be vaccinated because of age or health status to benefit from indirect protection from infection. Establishing effective public health policy including tailored national vaccination programs therefore requires an accurate understanding of immunity at a population level. Addressing this need requires broad access to quantitative serological testing, with an implicit requirement for scalable, non-invasive sample collection and high throughput testing. Saliva-based antibody testing has been reported as a non-invasive method for defining COVID-19 immunological status, and good correlation has been reported between salivary and blood-based antibody responses. However, these studies relied on the collection and testing of gingival crevicular fluid, which is known to be enriched in blood components, requires a specialized collection device and may be more challenging to scale.^9^ In the present study, we demonstrated the ability to measure protective IgG concentrations in whole saliva samples collected without specialized collection devices. We also demonstrated the ability to quantify protective IgG antibodies in DBS samples. Both saliva and DBS represent a path for collecting samples for immunity assessment remotely and non-invasively.

This study has certain limitations, the foremost being we examined only humoral (antibody-mediated) and not cellular-immunity; we hoped to provide initial support of the use of anti-spike IgG concentration as a correlate of protection for vaccine efficacy. Additional limitations include a small sample size of 37 donors total, and 13 donors used for saliva / DBS high-frequency temporal sampling. Conclusions about the kinetics of IgG increase and decrease were limited by our sampling frequency, which could not be done more frequently due to declining willingness of donors to provide finger-stick blood.

Challenges with vaccine access, combined with vaccine hesitancy in some communities continue to present barriers to achievement of herd immunity. When combined with a growing appreciation for limitations in durability of COVID-19 vaccine-induced immune response, an accurate understanding of population-based immunity becomes a critical input to inform public health policy including national vaccination programs, mask mandates, social distancing, and potentially more severe restrictions on mobility within or between communities. The work presented here establishes a potential pathway for non-invasive, broadly accessible collection of samples suitable for high-throughput quantitative serological testing to aid in defining SARS-CoV-2 immunity at population scale.

## Data Availability

All data produced in the present study are available upon reasonable request to the authors.

## Supplementary Information

**Supplementary Table 1.** Detailed donor information

| Cohort 1 ID | Cohort 2 ID | Gender | Vaccine | Matrix 1 | Matrix 2 | Prior Covid Infection |
| --- | --- | --- | --- | --- | --- | --- |
| - | D16 | Male | BNT162b2 | DBS | - | No |
| - | D17 | Female | BNT162b2 | DBS | - | No |
| - | D18 | Male | Ad26.COV2 | DBS | - | Yes, Antigen |
| - | D19 | Female | BNT162b2 | DBS | - | No |
| - | D20 | Male | mRNA-1273 | DBS | - | No |
| - | D21 | Female | mRNA-1273 | DBS | - | Yes, presumed |
| - | D22 | Male | mRNA-1273 | DBS | - | No |
| - | D23 | Female | BNT162b2 | DBS | - | No |
| - | D24 | Male | BNT162b2 | DBS | - | No |
| - | D25 | Male | BNT162b2 | DBS | - | No |
| - | D26 | Female | BNT162b2 | DBS | - | Yes, PCR |
| - | D27 | Male | mRNA-1273 | DBS | - | Yes, PCR |
| - | D28 | Male | mRNA-1273 | DBS | - | No |
| - | D29 | Male | BNT162b2 | DBS | - | No |
| - | D30 | Female | Ad26.COV2 | DBS | - | Yes, Antigen |
| - | D31 | Male | mRNA-1273 | DBS | - | No |
| - | D32 | Female | mRNA-1273 | DBS | - | No |
| - | D33 | Female | BNT162b2 | DBS | - | No |
| - | D34 | Female | BNT162b2 | DBS | - | No |
| - | D35 | Female | BNT162b2 | DBS | - | No |
| - | D36 | Female | mRNA-1273 | DBS | - | No |
| - | D37 | Male | mRNA-1273 | DBS | - | Yes, PCR |
| - | D38 | Male | Ad26.COV2 | DBS | - | No |
| - | D39 | Male | mRNA-1273 | DBS | - | No |
| D1 | - | Male | BNT162b2 | DBS | Saliva | No |
| D2 | - | Female | mRNA-1273 | DBS | Saliva | No |
| D3 | - | Female | BNT162b2 | DBS | Saliva | No |
| D4 | - | Male | BNT162b2 | DBS | Saliva | No |
| D5 | - | Female | BNT162b2 | DBS | Saliva | No |
| D6 | - | Male | mRNA-1273 | DBS | Saliva | No |
| D7 | - | Female | BNT162b2 | DBS | Saliva | No |
| D8 | - | Female | BNT162b2 | DBS | Saliva | Yes, PCR |
| D9 | - | Male | mRNA-1273 | DBS | Saliva | Yes, PCR |
| D10 | - | Female | mRNA-1273 | DBS | Saliva | No |
| D11 | - | Male | BNT162b2 | DBS | Saliva | No |
| D12 | - | Female | BNT162b2 | DBS | Saliva | No |
| D13 | - | Female | Ad26.COV2 | DBS | Saliva | No |
| D14 | - | Female | mRNA-1273 | - | Saliva | No |
| D15 | - | Female | BNT162b2 | - | Saliva | No |

**Supplementary Figure S1.**
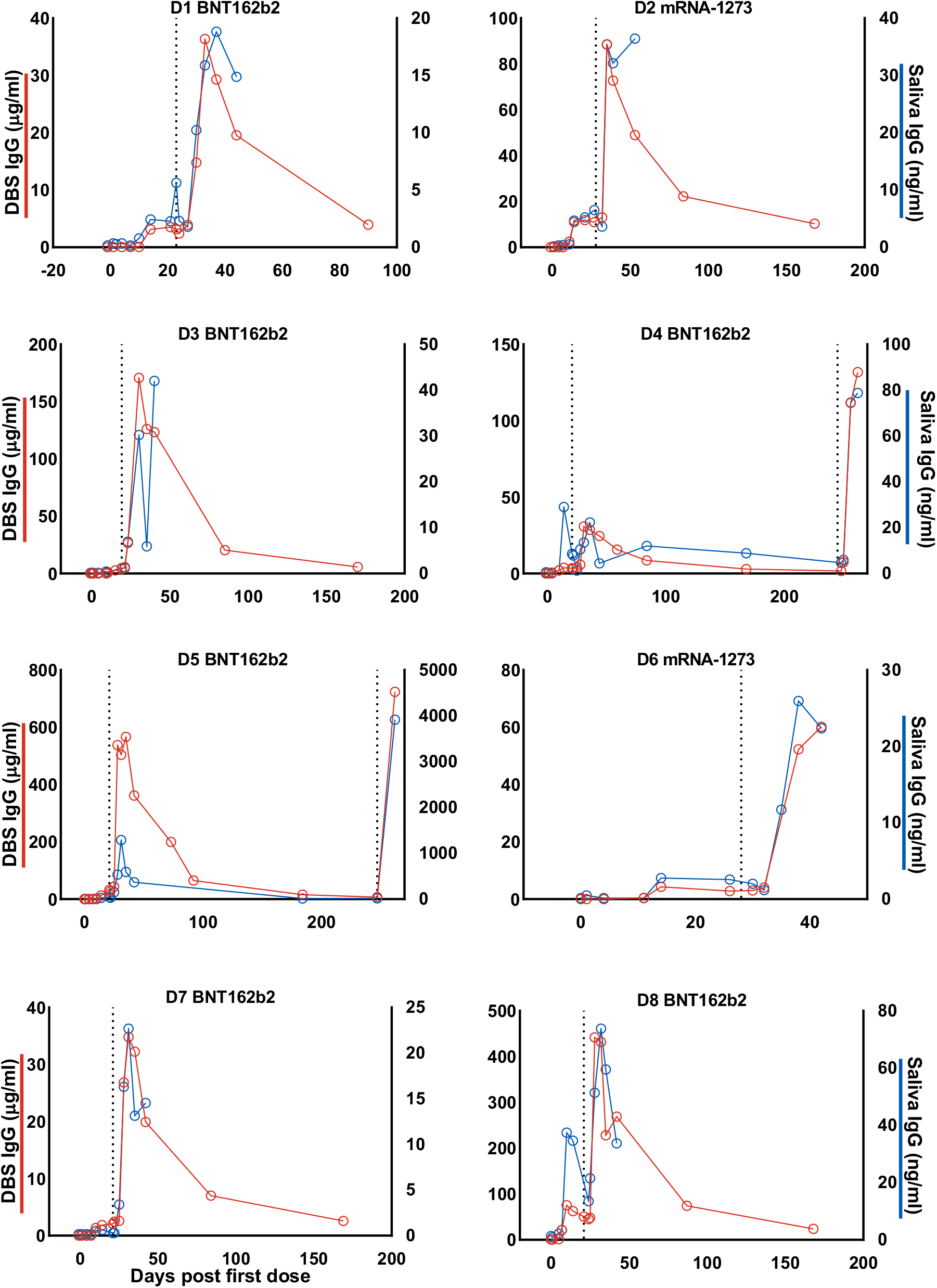
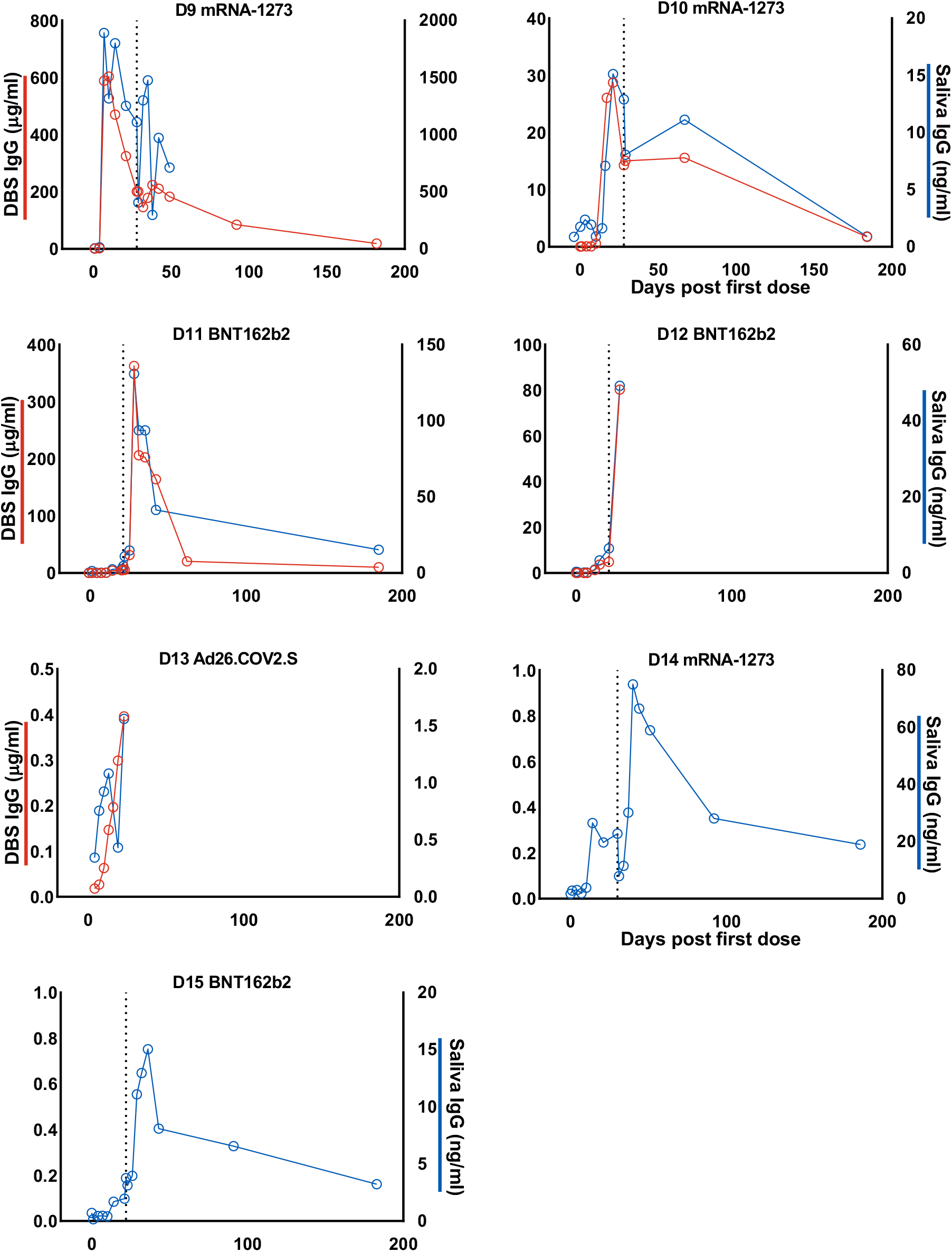
Longitudinal traces from individual donors of both DBS (red, left y-axis) and saliva (blue, right y-axis) before and after vaccination. The x-axis is numbered as days from first vaccination. The second day of 2^nd^ vaccination dose is shown as a dotted vertical line. Donor 122305 and 124387 provided samples after a third, booster dose. Donors 172610 and 190596 provided only saliva samples.

**Supplementary Figure S2.**
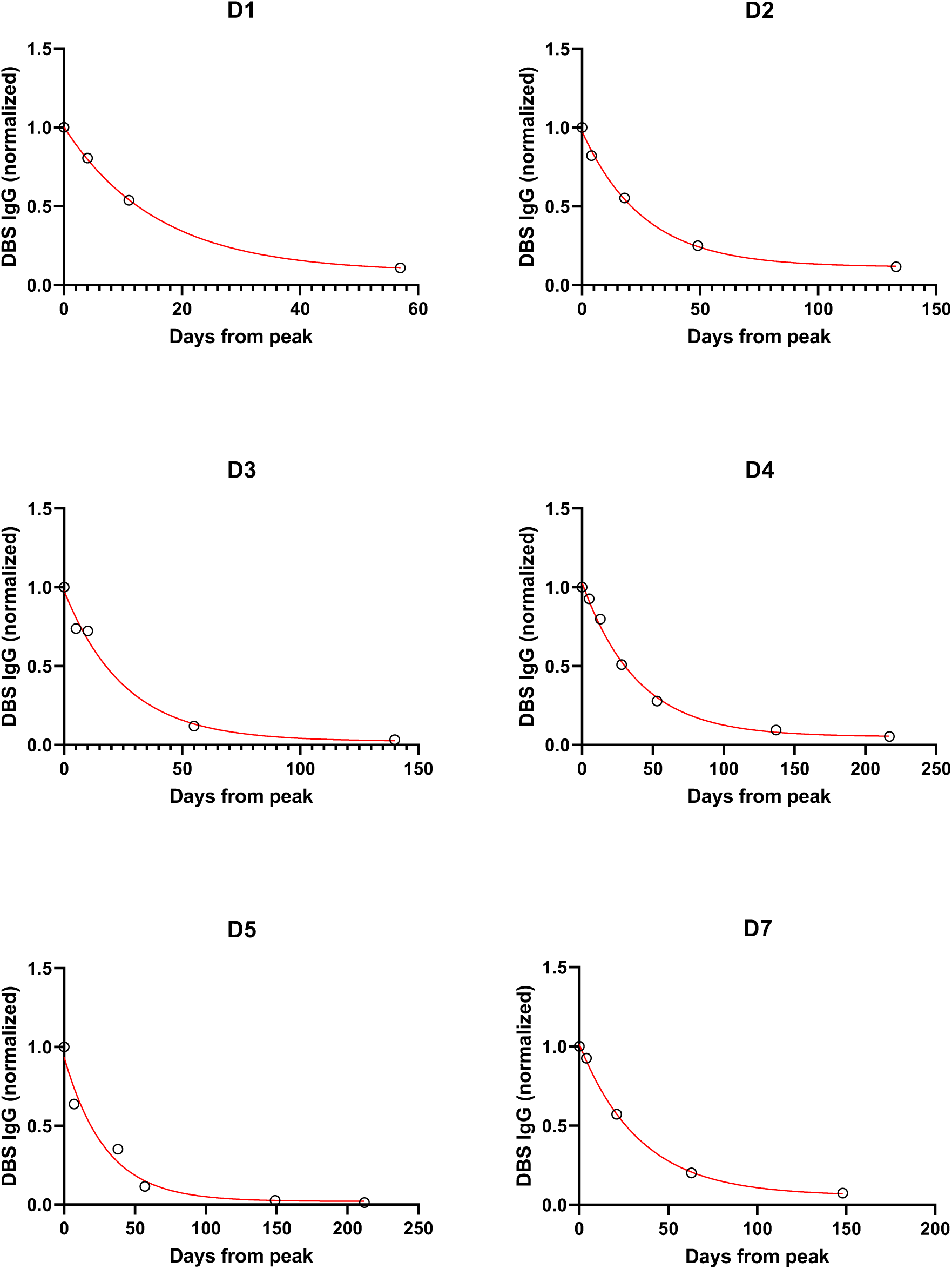
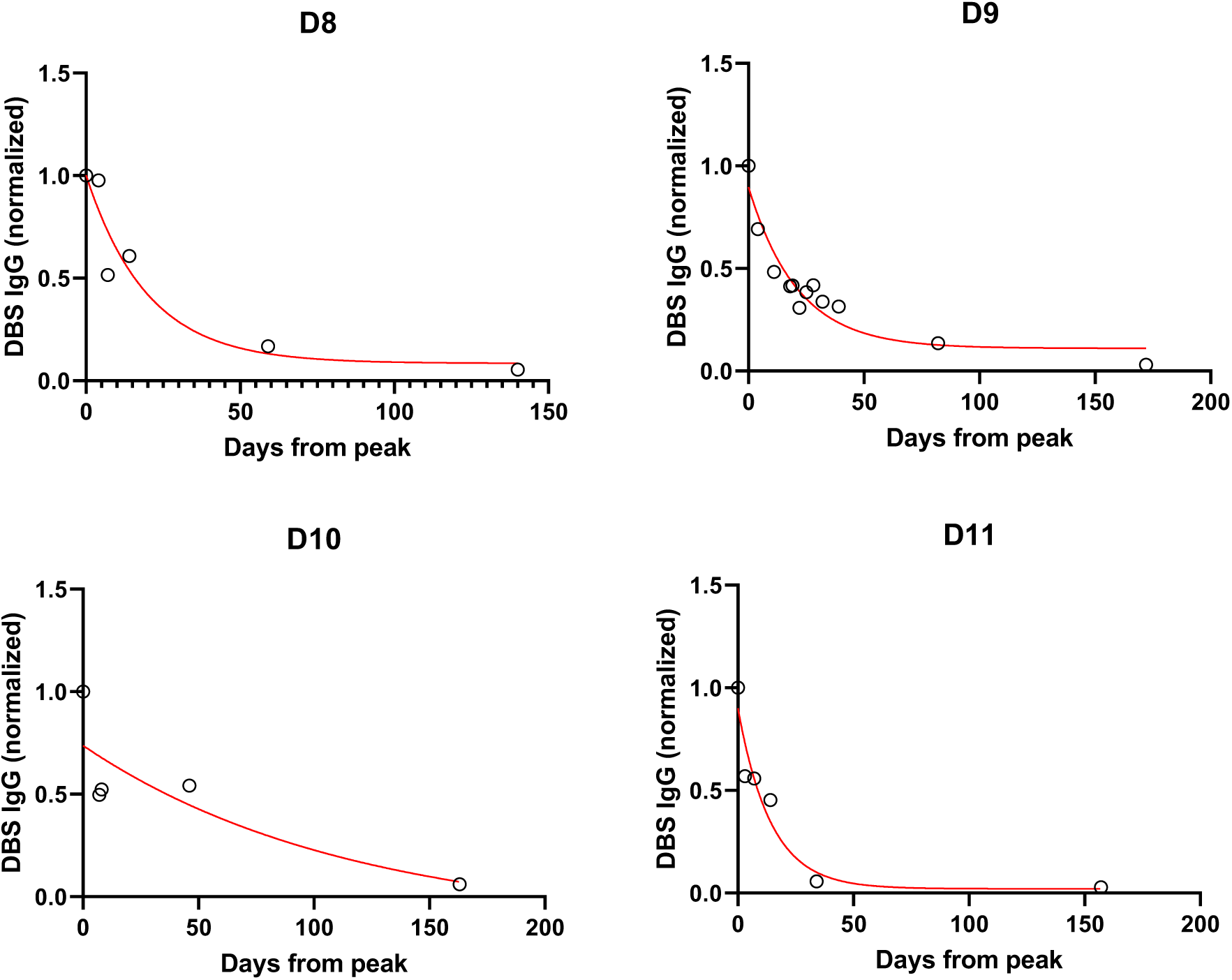
Decay of IgG levels in DBS, normalized to peak level. Longitudinal traces are shown from individual donors, along with one phase exponential decay regression (red lines).

**Supplementary Figure S3.**
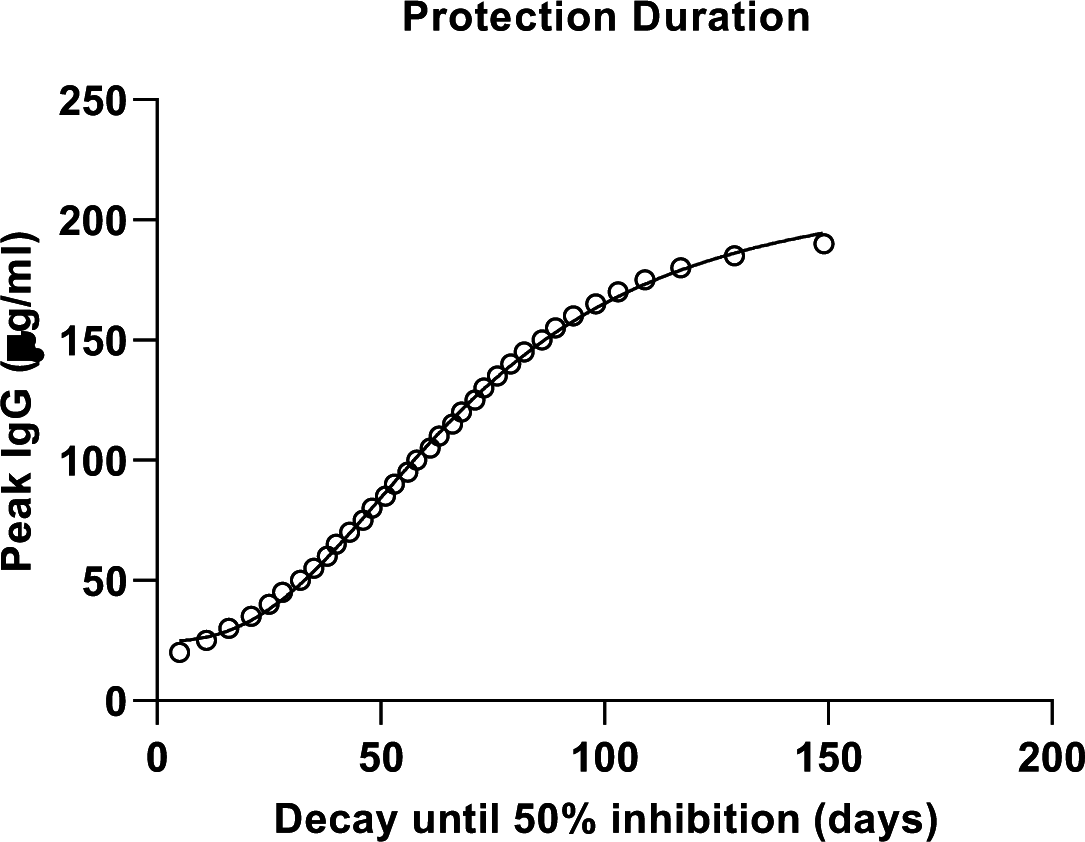
Duration of humoral protection based upon peak IgG levels in DBS. Based upon the exponential decay for all donors (Fig. 4e), the decay time to an IgG concentration of 15.7 ug/ml (corresponding to 50% binding-blocking) is predicted based upon peak IgG levels (circle symbols). The data fits well to a 4PL logistic regression with an R^2^=0.999.

## References

1. Jalkanen, P. et al. COVID-19 mRNA vaccine induced antibody responses against three SARS-CoV-2 variants. Nat. Commun. 2021 121 12, 1–11 (2021).

2. Wheeler, S. E. et al. Differential Antibody Response to mRNA COVID-19 Vaccines in Healthy Subjects. (2021) doi:10.1128/Spectrum.00341-21.

3. Khoury, D. S. et al. Neutralizing antibody levels are highly predictive of immune protection from symptomatic SARS-CoV-2 infection. Nat. Med. 27, 1205–1211 (2021).

4. Steensels, D., Pierlet, N., Penders, J., Mesotten, D. & Heylen, L. Comparison of SARS-CoV-2 Antibody Response Following Vaccination With BNT162b2 and mRNA-1273. JAMA 326, 1533–1535 (2021).

5. Boyarsky, B. J. et al. Antibody Response to 2-Dose SARS-CoV-2 mRNA Vaccine Series in Solid Organ Transplant Recipients. JAMA 325, 2204–2206 (2021).

6. Krammer, F. et al. Antibody Responses in Seropositive Persons after a Single Dose of SARS-CoV-2 mRNA Vaccine. N. Engl. J. Med. 384, 1372–1374 (2021).

7. Goel, R. R. et al. Distinct antibody and memory B cell responses in SARS-CoV-2 naïve and recovered individuals following mRNA vaccination. Sci. Immunol. 6, (2021).

8. Levin, E. G. et al. Waning Immune Humoral Response to BNT162b2 Covid-19 Vaccine over 6 Months. N. Engl. J. Med. (2021) doi:10.1056/NEJMoa2114583.

9. Pisanic, N. et al. COVID-19 Serology at Population Scale: SARS-CoV-2-Specific Antibody Responses in Saliva. J. Clin. Microbiol. 59, e02204–20 (2020).

10. Dobaño, C. et al. Persistence and baseline determinants of seropositivity and reinfection rates in health care workers up to 12.5 months after COVID-19. BMC Med. 19, 155 (2021).

11. Shan, D. et al. N-protein presents early in blood, dried blood and saliva during asymptomatic and symptomatic SARS-CoV-2 infection. Nat. Commun. 12, 1–8 (2021).

12. Rissin, D. M. et al. Single-molecule enzyme-linked immunosorbent assay detects serum proteins at subfemtomolar concentrations. Nat. Biotechnol. 28, 595–599 (2010).

13. Wilson, D. H. et al. The Simoa HD-1 Analyzer: A Novel Fully Automated Digital Immunoassay Analyzer with Single-Molecule Sensitivity and Multiplexing. J. Lab. Autom. 21, 533–547 (2016).

14. Kan, C. W. et al. Isolation and detection of single molecules on paramagnetic beads using sequential fluid flows in microfabricated polymer array assemblies. Lab Chip 12, 977–985 (2012).

15. Robin, X. et al. pROC: an open-source package for R and S+ to analyze and compare ROC curves. BMC Bioinformatics 12, 77 (2011).

16. Bakdash, J. Z. & Marusich, L. R. Repeated Measures Correlation. Front. Psychol. 8, 456 (2017).

17. de la Monte, S. M. et al. Heterogeneous Longitudinal Antibody Responses to Covid-19 mRNA Vaccination. Clin. Pathol. (Thousand Oaks, Ventur. County, Calif.) 14, 2632010X211049255 (2021).

18. Wei, J. et al. Antibody responses to SARS-CoV-2 vaccines in 45,965 adults from the general population of the United Kingdom. Nat. Microbiol. 6, 1140–1149 (2021).

19. Kongsuphol, P. et al. A rapid simple point-of-care assay for the detection of SARS-CoV-2 neutralizing antibodies. Commun. Med. 1, 46 (2021).

20. U.S. Center for Disease Control. COVID-19 Vaccine Booster Shots. https://www.cdc.gov/coronavirus/2019-ncov/vaccines/booster-shot.html?s_cid=11706:cdccovidbooster:sem.ga:p:RG:GM:gen:PTN:FY22 (2021).

21. Anderson, M. et al. SARS-CoV-2 Antibody Responses in Infection-Naive or Previously Infected Individuals After 1 and 2 Doses of the BNT162b2 Vaccine. JAMA Netw. open 4, e2119741–e2119741 (2021).

22. Bradley, T., Grundberg, E. & Selvarangan, R. Antibody responses boosted in seropositive healthcare workers after single dose of SARS-CoV-2 mRNA vaccine. medRxiv : the preprint server for health sciences (2021) doi:10.1101/2021.02.03.21251078.

23. Ebinger, J. E. et al. Prior COVID-19 Infection and Antibody Response to Single Versus Double Dose mRNA SARS-CoV-2 Vaccination. medRxiv : the preprint server for health sciences (2021) doi:10.1101/2021.02.23.21252230.

24. Collier, A. Y. et al. Differential Kinetics of Immune Responses Elicited by Covid-19 Vaccines. N. Engl. J. Med. 385, 2010–2012 (2021).

25. Gobbi, F. et al. Antibody Response to the BNT162b2 mRNA COVID-19 Vaccine in Subjects with Prior SARS-CoV-2 Infection. Viruses 13, 422 (2021).

26. Canaday, D. H. et al. Significant reduction in humoral immunity among healthcare workers and nursing home residents 6 months after COVID-19 BNT162b2 mRNA vaccination. medRxiv 2021.08.15.21262067 (2021) doi:10.1101/2021.08.15.21262067.

27. Marot, S. et al. Rapid decline of neutralizing antibodies against SARS-CoV-2 among infected healthcare workers. Nat. Commun. 12, 2824 (2021).

28. Xiang, T. et al. Declining Levels of Neutralizing Antibodies Against SARS-CoV-2 in Convalescent COVID-19 Patients One Year Post Symptom Onset. Front. Immunol. 12, 708523 (2021).

29. Wang, Z. et al. mRNA vaccine-elicited antibodies to SARS-CoV-2 and circulating variants. Nature 592, 616–622 (2021).

30. Jiang, S., Zhang, X., Yang, Y., Hotez, P. J. & Du, L. Neutralizing antibodies for the treatment of COVID-19. Nat. Biomed. Eng. 4, 1134–1139 (2020).

31. Ketas, T. J. et al. Antibody responses to SARS-CoV-2 mRNA vaccines are detectable in saliva. bioRxiv : the preprint server for biology (2021) doi:10.1101/2021.03.11.434841.

32. Sheikh-Mohamed, S. et al. A mucosal antibody response is induced by intra-muscular SARS-CoV-2 mRNA vaccination. medRxiv 2021.08.01.21261297 (2021).

33. Brandtzaeg, P. Secretory immunity with special reference to the oral cavity. J. Oral Microbiol. 5, (2013).

34. Heinzel, C. et al. Non-Invasive Antibody Assessment in Saliva to Determine SARS-CoV-2 Exposure in Young Children. Front. Immunol. 12, 753435 (2021).

35. Alkharaan, H. et al. Persisting salivary IgG against SARS-CoV-2 at 9 months after mild COVID-19: A complementary approach to population surveys. J. Infect. Dis. 224, 407–414 (2021).

36. MacMullan, M. A. et al. ELISA detection of SARS-CoV-2 antibodies in saliva. Sci. Rep. 10, 20818 (2020).

37. Anderson, R. M. & May, R. M. Vaccination and herd immunity to infectious diseases. Nature 318, 323–329 (1985).

